# Structural heterogeneity reveals distinct amyloidopathies in Alzheimer’s disease

**DOI:** 10.1101/2025.11.17.25340406

**Authors:** Jan Sebastian Novotný, Mária Čarná, Iain Lyburn, Tarun Kuruvilla, Gorazd Bernard Stokin, the Alzheimer’s Disease Neuroimaging Initiative

## Abstract

Within the amyloid/tau/neurodegeneration (ATN) framework, hippocampal (H) and inferior parietal lobule (IPL) atrophy are established markers of neurodegeneration (N+), whereas the biological significance of choroid plexus (ChP) enlargement remains uncertain. Although amyloid positivity (A+) underpins the current biological definition of Alzheimer’s disease, the extent to which it is consistently associated with structural neurodegeneration remains unclear. We investigated the relationship between A+ and structural neurodegeneration using structural MRI, amyloid PET, and cognitive data from 872 participants from the Alzheimer’s Disease Neuroimaging Initiative (ADNI). Regional brain volumes (H, IPL and ChP) were classified as abnormal using validated normative thresholds. Diagnostic performance was evaluated using multiple machine-learning approaches, and associations between structural phenotypes, cognitive profiles and cerebrospinal fluid (CSF) biomarkers were examined.

Among 872 participants, 411 were amyloid-negative healthy subjects, 339 had amyloid-positive mild cognitive impairment, and 122 had amyloid-positive Alzheimer’s disease dementia. At the group level, H and IPL progressively decreased, whereas ChP volume increased across disease stages (all P<0.001). However, 45% of amyloid-positive individuals exhibited no detectable structural neurodegeneration. Consistently, machine-learning models demonstrated limited individual-level diagnostic performance, with a maximum overall accuracy of 0.60 [0.57–0.62]. Hierarchical clustering identified four cognitive phenotypes that mapped onto distinct structural signatures.

Amyloid positivity alone does not uniformly predict structural neurodegeneration. The marked dissociation between amyloid pathology and MRI-defined neurodegeneration demonstrates substantial biological heterogeneity within amyloid-positive Alzheimer’s disease and supports the concept of distinct structural amyloidopathies. These findings have important implications for biomarker interpretation, patient stratification, and the development of precision therapeutic strategies.

## Introduction

One of the central assumptions underlying the biological definition of Alzheimer’s disease (AD) is that amyloid pathology ultimately leads to neurodegeneration. This concept forms the foundation of the amyloid cascade hypothesis [1, 2], underpins the National Institute on Aging–Alzheimer’s Association (NIA-AA) ATN biological framework [3], and provides the rationale for the recent development of amyloid-targeting therapies [4, 5]. Yet despite its central importance, the extent to which amyloid positivity (A+) consistently translates into structural neurodegeneration (N+) at the individual level remains uncertain. Increasing reports of discordance between amyloid biomarkers and neurodegeneration challenge the concept of a uniform amyloid-driven neurodegenerative cascade and suggest substantially greater biological heterogeneity than predicted by linear models of disease progression [6–8]. As disease-modifying anti-amyloid therapies enter routine clinical practice, understanding this relationship has become an increasingly important biological and clinical question.

Structural MRI markers of neurodegeneration, particularly hippocampal (H) and inferior parietal lobule (IPL) atrophy, constitute the principal imaging measures of N+ within the ATN framework [3, 9]. These markers are embedded in contemporary diagnostic criteria [10, 11] and have been widely used to define imaging-based subtypes of Alzheimer’s disease [12, 13]. However, tau pathology frequently demonstrates stronger associations with neuronal loss and clinical severity than amyloid pathology itself [14, 15], raising the possibility that amyloid positivity alone may not fully determine downstream structural degeneration. Consequently, the biological coupling between A+ and MRI-defined neurodegeneration remains incompletely understood.

Among emerging structural biomarkers, enlargement of the choroid plexus (ChP) has attracted increasing interest. Multiple imaging studies have reported increased ChP volume in Alzheimer’s disease and mild cognitive impairment [16–20], potentially reflecting alterations in cerebrospinal fluid dynamics, glymphatic clearance, blood-brain barrier integrity, neurovascular dysfunction, or neuroinflammatory processes. Unlike hippocampal and cortical atrophy, however, the biological significance of ChP enlargement and its relationship to established ATN biomarkers remain poorly defined. Whether ChP enlargement represents an independent pathological process or an integral component of Alzheimer’s disease progression is therefore unknown.

Here, we investigated whether amyloid positivity is consistently associated with structural neurodegeneration across the Alzheimer’s disease spectrum. Using structural MRI, amyloid PET, cognitive assessment, cerebrospinal fluid biomarkers, and machine-learning approaches in 872 participants from the Alzheimer’s Disease Neuroimaging Initiative (ADNI), we evaluated the diagnostic utility of established neurodegeneration markers (H and IPL), examined the contribution of ChP enlargement, and explored the relationship between structural phenotypes, cognition, and molecular biomarkers.

Our findings reveal a striking dissociation between amyloid positivity and structural neurodegeneration at the individual level. Nearly half of amyloid-positive participants exhibited no detectable MRI-defined neurodegeneration despite biomarker-confirmed amyloid pathology, while distinct structural phenotypes corresponded to divergent cognitive profiles and biomarker associations. These observations indicate that amyloid positivity alone does not uniformly determine downstream structural neurodegeneration and support substantial biological heterogeneity within amyloid-positive Alzheimer’s disease. Rather than representing a single stereotyped neurodegenerative process, our findings are consistent with the existence of distinct structural amyloidopathies, with important implications for biomarker interpretation, patient stratification, and the development of precision therapeutic strategies.

## Methods

### Study Overview

This retrospective observational study analysed structural MRI, amyloid PET, cognitive, genetic and cerebrospinal fluid biomarker data from participants enrolled in the Alzheimer’s Disease Neuroimaging Initiative (ADNI). The primary objective was to determine whether amyloid positivity consistently predicts structural neurodegeneration at the individual level and to identify structural phenotypes associated with cognitive and biomarker profiles.

### Study Design and Data Source

Data were obtained from the ADNI database, for up-to-date information, see www.adni-info.org. Launched in 2003 as a public-private partnership, led by Principal Investigator Michael W. Weiner, ADNI aims to determine whether serial magnetic resonance imaging (MRI), positron emission tomography (PET), biological markers, and clinical and neuropsychological assessments can be integrated to track progression of mild cognitive impairment (MCI) and early Alzheimer’s disease (AD).

### Participants and Clinical Classification

Participants were included if baseline MRI, amyloid PET and standardized cognitive assessments – comprising the Mini-Mental State Examination (MMSE), ADNI Cognitive Composite Score (ADNI CogSum), and Clinical Dementia Rating [CDR]) – were included.

The final sample comprised 872 individuals (mean age 71.8±7.0 years; 444 women [50.9%]). Based on clinical and biomarker profiles, participants were stratified into three diagnostic groups: healthy subjects (HS; n=411): A^-^ PET, CDR = 0, and MMSE ≥ 24 (mean age=70.2±6.6), mild cognitive impairment due to AD (MCI; n=339): A^+^ PET, CDR = 0.5, and MMSE > 24 (mean age=72.8±6.8), and AD dementia (D; n=122): A^+^ PET, CDR > 1, and MMSE score < 24 (mean age=74.2±7.9) (Figure 1a, Supplementary Table S1).

**Figure 1.**
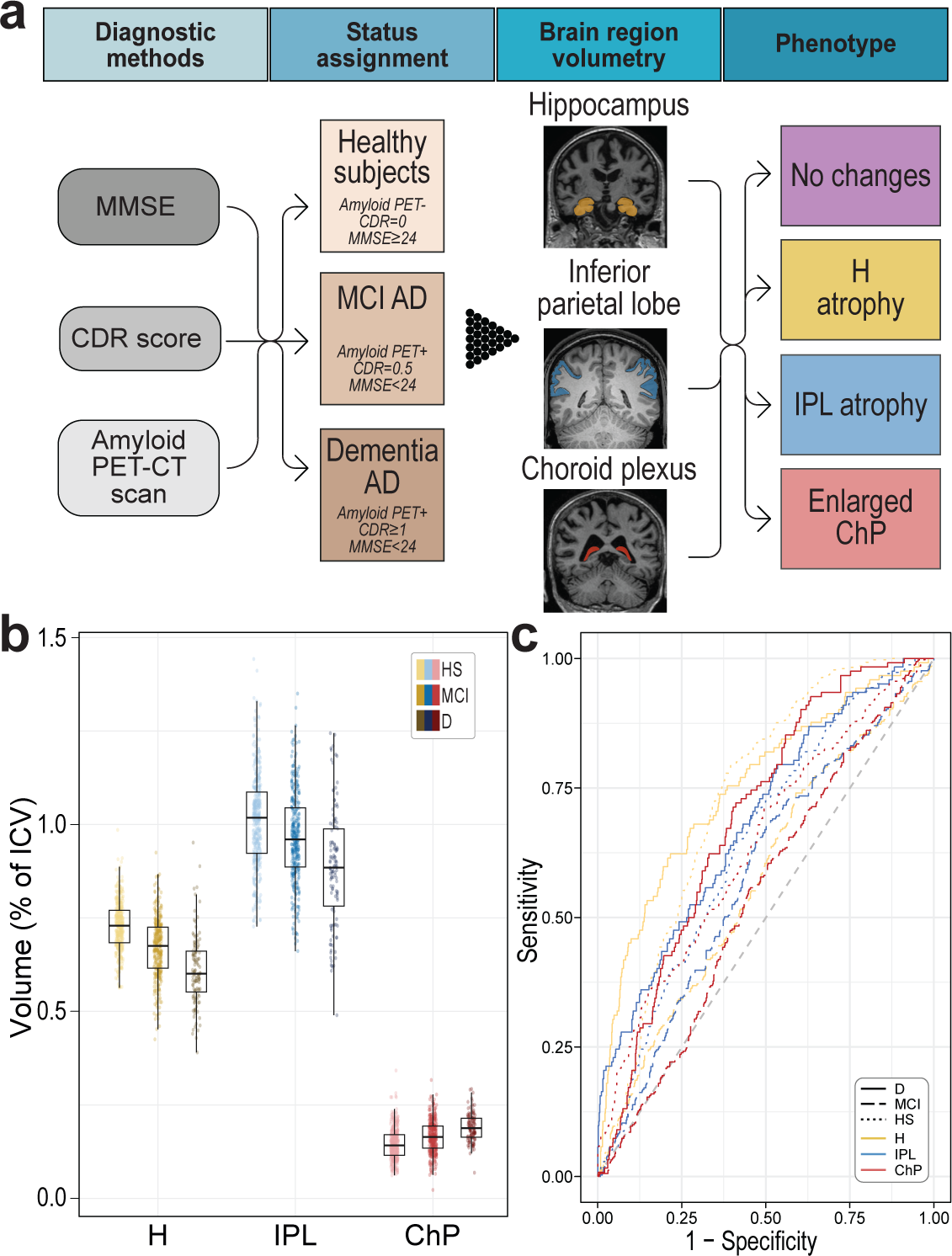
Association between structural brain changes (*N*) and diagnostic status. (a) Schematic of the study pipeline for diagnostic classification and derivation of volumetric phenotypes. (b) Distribution of normalized hippocampal, inferior parietal lobule, and choroid plexus volumes across healthy controls (*A*^-^) and amyloid-positive (*A*^+^) mild cognitive impairment and dementia participants. (c) Receiver operating characteristic (ROC) curves of Artificial Neural Network multiclass predictions of diagnostic status based on regional volumes (H, IPL, and ChP). **Abbreviations**: HS, healthy subjects; MCI, mild cognitive impairment; D, AD dementia; H, hippocampus; IPL, inferior parietal lobule; ChP, choroid plexus.

APOE ε4 genotype data were available for 885 participants (98.0%), and cerebrospinal fluid (CSF) biomarker data (Aβ_1-42_, total tau, phosphorylated tau [p-tau181]) for 159 participants (18.2%).

### MRI Acquisition and Neurodegeneration (N) Processing

Regional brain volumes were derived from standardised ADNI pipelines (UCSF UCSFFSX7 and UCSFFSL51 datasets). T1-weighted images were processed using FreeSurfer (v7.x cross-sectional, v5.1 longitudinal) to ensure anatomical consistency [21, 22]. Based on our study hypothesis, analyses focused on three primary regions of interest (ROIs): the hippocampus (H) and inferior parietal lobule (IPL), established markers of neurodegeneration (*N*^+^), and the choroid plexus (ChP) was evaluated as an exploratory structural biomarker. ROI volumes were summed across hemispheres and normalized to total intracranial volume (TIV) using a regression-based approach to account for inter-individual variability in head size [23, 24].

For participants with longitudinal MRI data, a representative scan was selected by computing the mean of normalized ROI volumes across timepoints (using reciprocally transformed values for ChP) and identifying the scan closest to this participant-specific mean. This approach was implemented to minimize measurement noise and ensure that structural measures reflected stable neuroanatomical state.

### Biomarker Definitions and ATN Status

Amyloid (*A*) status was determined via PET using validated pipelines harmonized across tracers (^11^C-PiB, ^18^F-florbetapir, ^18^F-florbetaben, and ^18^F-NAV4694)[25]. PET scans were acquired within two weeks of baseline clinical assessment [26], and *A*^+^ status was defined using established UC Berkeley thresholds [25, 27].

Tau (*T*) status was determined via CSF concentrations of total tau and p-tau 181 using ADNI-specific thresholds.

Neurodegeneration (*N*) was defined as H or IPL atrophy falling below the 5th percentile of the normative HS distribution [28]. ChP enlargement was defined as volume exceeding the 90th percentile of the normative distribution. This conservative threshold was selected to identify significant enlargement while minimising false positives in the absence of established clinical cut-offs.

### Laboratory and Genotype Analysis

APOE-ε4 genotyping was performed using DNA extracted from EDTA blood samples collected at screening [29]. CSF samples were obtained via lumbar puncture, processed within one hour, and stored at −80°C. Biomarker concentrations (Aβ_1–42_, total-tau, and p-tau181]) were measured using Elecsys immunoassays (cobas e601 analyzer, University of Pennsylvania Biomarker Core) [30].

### Statistical analysis

Analyses were conducted on complete cases. Group differences in regional volumes (*N* markers) were assessed using analysis of covariance (ANCOVA), adjusting for age, sex, and education. Associations between structural biomarkers (*N^+^*), biomarker status (*A*^+^, *T*^+^), and diagnostic category were evaluated using multinomial logistic regression (MLR).

To assess the robustness of diagnostic classification, predictive performance was evaluated using three complementary machine-learning algorithms representing linear (Multinomial Logistic Regression), ensemble (Random Forest), and nonlinear (Artificial Neural Network) approaches. Model robustness was ensured via 5-fold cross-validation. Categorical volumetric patterns (atrophy, enlargement, and their combinations) and sex-specific models were further evaluated using ANN.

Differences in the prevalence of volumetric patterns and APOE ε4 distribution were assessed using Chi-square tests (Cramer’s V reported for effect size). Cognitive differences were analysed via analysis of variance (ANOVA) with Dunnett’s post-hoc comparisons against HS and the “no atrophy” groups. Hierarchical cluster analysis (Ward’s method) was used to classify cognitive phenotypes.

All tests were two-tailed (p<0.05), with multiple comparisons controlled via the Benjamini-Hochberg procedure. Statistical analyses were performed in RStudio (v.2022.07.2; R version 4.2.1).

### Sex Stratified and Interaction Analyses

To examine the stability of findings across sexes, brain volumes were compared separately within each sex using ANCOVA. A diagnostic group × sex interaction term was included to test for sexual dimorphism. Predictive accuracies between sexes were compared using the DeLong test. Finally, the associations between cognitive phenotypes and volumetric patterns were examined using the Cochran-Mantel-Haenszel test, stratified by sex. Sex differences in CSF biomarker levels according to atrophy or enlargement were assessed using linear models.

### Reproducibility and Transparency

All data used in this study are publicly available through the ADNI database (www.adni.loni.usc.edu). Imaging data were processed using standardized and widely used pipelines, and all preprocessing stems are documented within ADNI. Statistical analyses were performed using established methods and open-source software (R). Thresholds for volumetric classification were defined a priori based on normative distributions. No outcome-driven threshold optimisation was performed. No data imputation was performed; analyses were based on complete cases.

## Results

### Structural Changes Across the Diagnostic Spectrum

Group-level structural neurodegeneration (*N*^+^) followed the expected pattern across the Alzheimer’s disease spectrum, but substantial heterogeneity emerged at the individual level. A progressive reduction in H/IPL volumes was observed across diagnostic groups (H volume: HS 0.73±0.07 vs. MCI 0.67±0.08 vs. D 0.61±0.09; F[1,2]=11.2; P<0.001; partial η²=0.21), accompanied by a reciprocal increase in ChP volume (HS 0.15±0.04 vs. MCI 0.16±0.04 vs. D 0.19±0.04; F[1,2]=52.44; P<0.001; partial η²=0.09; Figure 1b, Supplementary Figure 1, Supplementary Table S2-3). Despite these robust group-level trends, machine learning (ML) models failed to reliably discriminate between *A*^-^ healthy subjects and *A*^+^ clinical cases; the artificial neural network combined metrics (H+IPL+ChP) yielded a peak overall accuracy of only 0.60 [95% CI 0.57-0.62], with a sensitivity of 0.76 for HS, 0.49 for MCI, and 0.35 for D (Cohen’s κ=0.31±0.06; Figure 1c, Supplementary Figures 2-3, Supplementary Table S4). Diagnostic classification was substantially hampered by marked inter-individual variability within each clinical group (Supplementary Figure 4). Additionally, APOE ε4 status was not significantly associated with any structural *N* measure (all p>0.383; Supplementary Figure 5, Supplementary Table S5).

### Prevalence and Longitudinal Dynamics of N Abnormalities

The prevalence of structural abnormalities varied significantly across *A*^+^ groups. In the *A*^+^ MCI group, H atrophy, IPL atrophy, and ChP enlargement were present in only 27.7%, 9.1%, and 21.5% of participants, respectively (all P<0.001 vs. dementia group; Cramér’s V range, 0,18-0.27). In the *A*^+^ dementia group, these proportions increased to 57.4%, 27.9%, and 40.2% (prevalence ratio vs. MCI: H 2.07 [95% CI 1.64-2.60]; IPL 3.05 [1.96-4.74]; ChP 1.87 [1.39-2.51]; Figure 2a, Supplementary Table S6). In the present study, “N^-^” refers specifically to the absence of detectable macroscopic structural abnormalities on volumetric MRI and should not be interpreted as absence of molecular, synaptic, or microstructural neurodegeneration. Notably, nearly half (45%) of all *A*^+^ individuals exhibited no volumetric MRI changes. Conversely, isolated H atrophy, IPL atrophy, and ChP enlargement were observed in 19%, 5%, and 12% of cases, respectively (Figure 2b).

**Figure 2.**
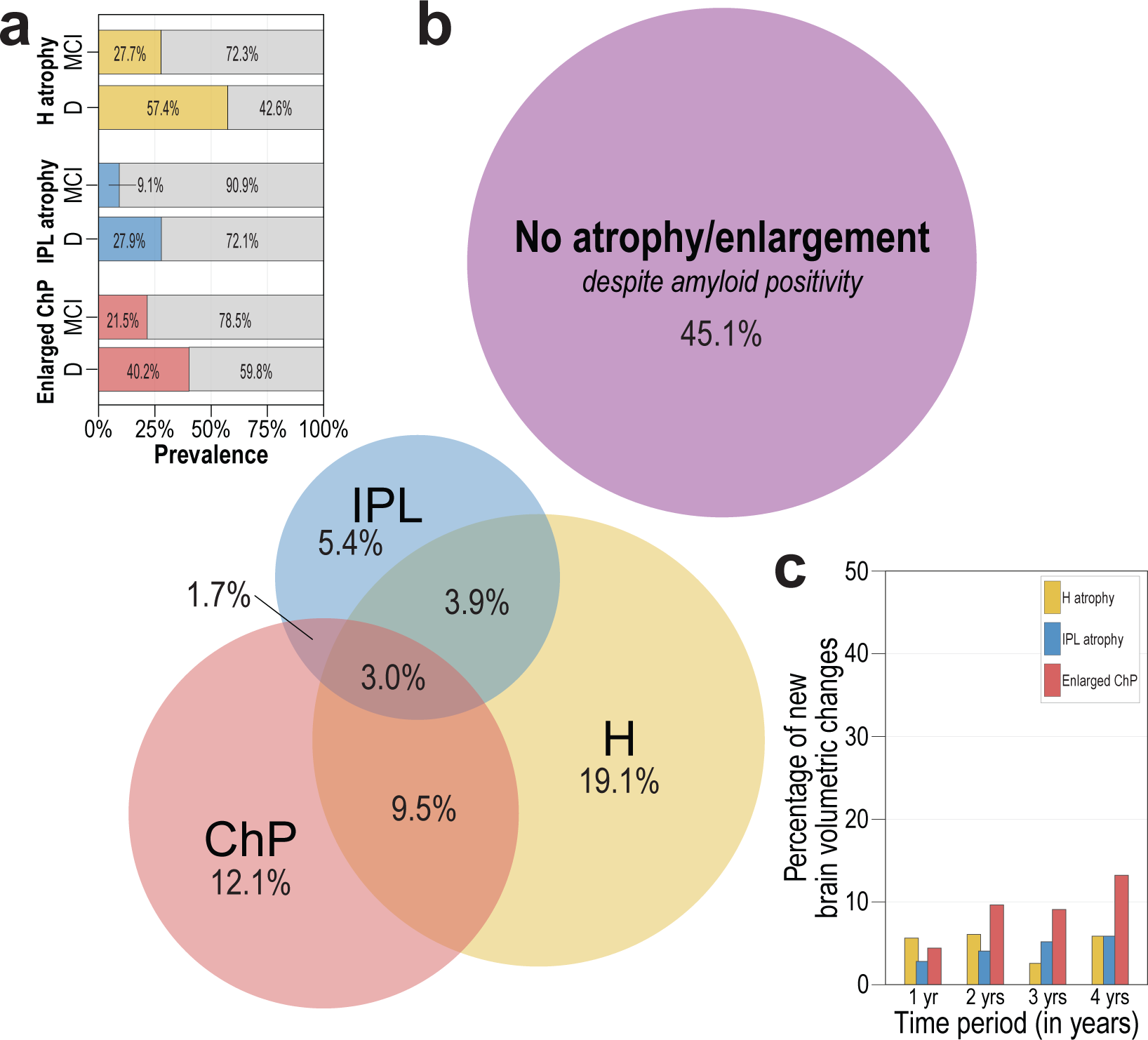
Prevalence and longitudinal dynamics of structural neurodegeneration (*N*) in Aβ^+^ individuals. (a) Prevalence of H atrophy, IPL atrophy, and ChP enlargement in A^+^ MCI and A^+^ dementia cohorts. (b) Venn diagram illustrating the overlap between regional structural abnormalities and the proportion of *A*^+^ cases exhibiting no detectable structural abnormalities (*N*^-^). (c) Longitudinal trajectories of structural change over a 4-year follow-up. Bars represent the incidence of new structural abnormalities per interval; note that the vast majority of participants (∼ 85-95%) remained stable. **Abbreviations**: HS, healthy subjects; MCI, mild cognitive impairment; D, dementia.

Over four years of longitudinal follow-up, rates of H and IPL atrophy, as well as ChP enlargement, remained relatively stable (H: Z=0, P>0.99, Sen’s slope=-0.01, IPL: Z=1.698, P=0.089, Sen’s slope=1.08, ChP: Z=1.019, P=0.308, Sen’s slope=2.63; Figure 2c, Supplementary Table S7). Predictive models based on categorical *N* phenotypes likewise showed poor diagnostic classification performance with best overall accuracy of 0.56 [95% CI 0.55-0.57] observed in model using H+IPL, sensitivity of 0.93 for HS, 0.25 for MCI, and 0.17 for D (Cohen κ=0.21±0.02) and overall accuracy of 0.55 [95% CI 0.52-0.58] for full H+IPL+ChP model, sensitivity of 0.84 for HS, 0.33 for MCI, D 0.17 for D (Cohen κ=0.20±0.04; Supplementary Figure 6, Supplementary Table S8).

### Cognitive Phenotypes and Structural Mapping

Global cognitive performance (MMSE) was significantly lower in both *A*^+^ MCI and *A*^+^ dementia compared to *A*^-^ healthy subjects, yet it was only weakly associated with structural *N* status (Supplementary Figure 7a, Supplementary Table S9). The greatest impairment was observed in individuals with concurrent H and IPL atrophy, and ChP enlargement. Similar patterns of dissociation were observed across domain-specific cognitive measures derived from ADNI CogSum scores (Supplementary Figure 7b-e, Supplementary Table S10).

Hierarchical clustering identified four distinct cognitive phenotypes that mapped closely to specific structural patterns (Figure 3a, Supplementary Table S11). Cluster 1 (severe global impairment; mean MMSE 16.3±5.4) was enriched for multi-domain structural abnormalities (volumetric changes in H, IPL, and ChP). Cluster 4 (minimal impairment; mean MMSE 28.2±1.6) corresponded to a complete absence of structural N (*N^-^*). Intermediate clusters were characterized by specific regional signatures: Cluster 2 was enriched for isolated H atrophy, whereas Cluster 3 was primarily associated with isolated ChP enlargement (Figure 3b, Supplementary Table S12).

**Figure 3.**
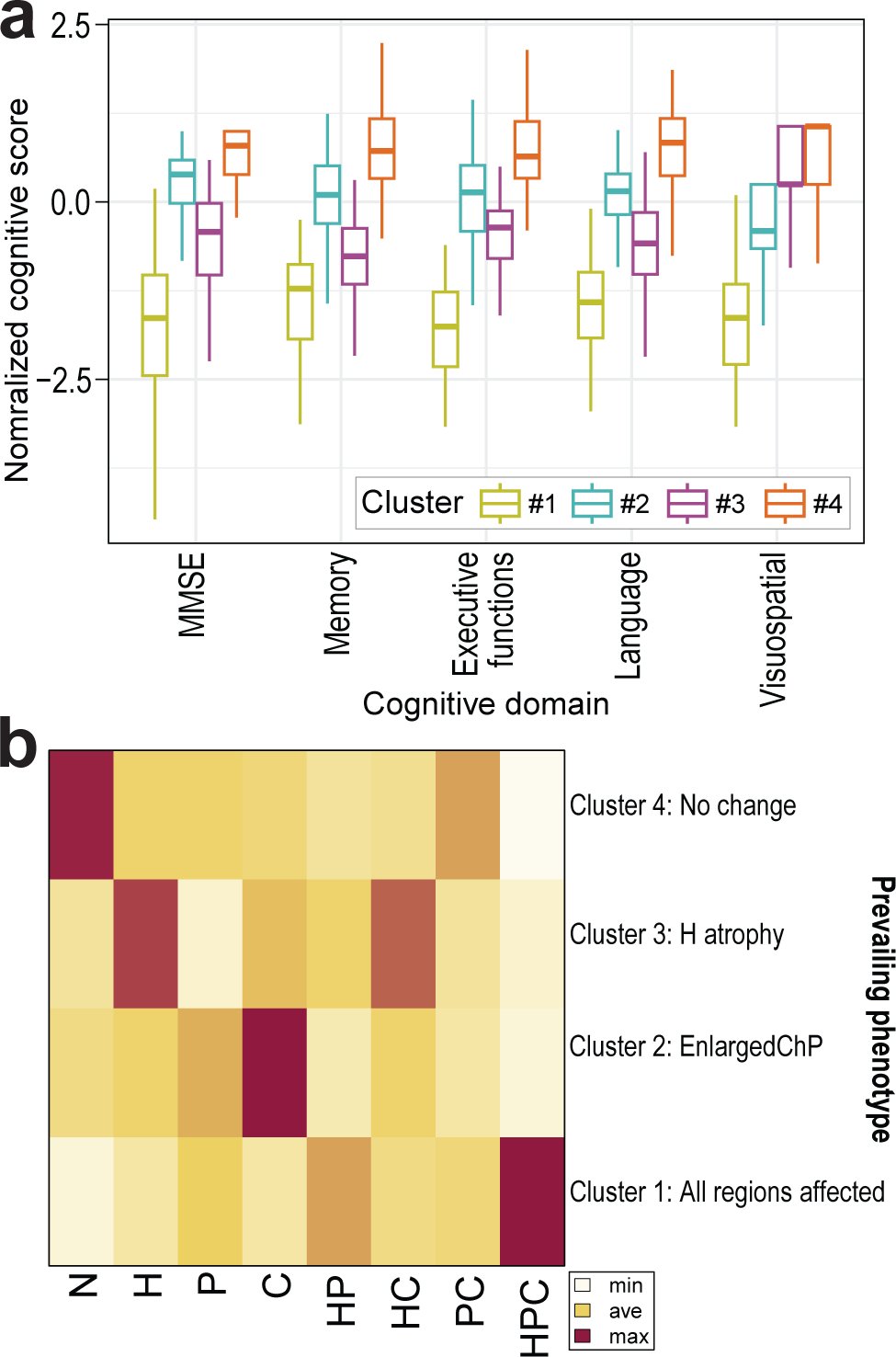
Cognitive profiles in relation to structural phenotypes. (a) Scaled Mini-Mental State Examination (MMSE) and ADNI CogSum domain scores across four cognitive clusters identified via hierarchical clustering. (b) Distribution of volumetric phenotypes within each cognitive cluster (row-wise percentages). **Abbreviations**: N, no volumetric change, H: hippocampal atrophy, P: inferior parietal lobule atrophy, C: choroid plexus enlargement, HP: hippocampal and inferior parietal lobule atrophy, HC: hippocampal atrophy with choroid plexus enlargement, HPC: hippocampal and inferior parietal lobule atrophy with choroid plexus enlargement.

### Associations with CSF Biomarkers

We examined associations between CSF biomarkers (*A* and *T*) and structural *N* measures. While H and IPL atrophy were associated with lower CSF Aβ_1-42_ levels (H atrophy: 585±165 vs. 773±299 pg/mL; Cohen’s d=0.72; P=0.005; IPL atrophy: 573±152 vs. 737±287 pg/mL; d=0.61; P=0.022), consistent with higher amyloid plaque burden, ChP enlargement showed no such association (752±368 vs. 701±256 pg/mL; P=0.696). Crucially, CSF total and phosphorylated tau (*T*+) levels did not differ significantly based on structural N status, regardless of the brain region affected (Supplementary Figure 8, Supplementary Table S13).

### Inter-Sex Stability of Findings

To assess the stability of findings across sexes, we performed sex-specific analyses. Females exhibited marginally higher H volumes in the HS group and lower IPL volumes in the D group (Supplementary Figure 9, Supplementary Table S14). Consistent with the overall sample, nearly half of *A^+^* participants exhibited no structural abnormalities (females: 48%; males: 42.5%; Supplementary Figure 10).

Although diagnostic prediction based on volumetric data was modestly improved in females – particularly when H volume was included ((AUC for HS: 0.791 in females vs. 0.696 in males; DeLong P=0.004; combined H+IPL+ChP: AUC 0.803 vs. 0.724; P=0.014; Supplementary Table S15) – overall classification accuracy remained poor for both sexes. Detection rates for HS were below 50% (females ∼83–85%; males ∼70–75%) and generally below 20% and 35% for MCI and D in both sexes, respectively (Supplementary Figure 11, Supplementary Table S16). Predictions based on categorical *N* phenotypes did not differ by sexes (Supplementary Table S17), and accuracy remained similarly inadequate (Supplementary Figure 12, Supplementary Table S18). Structural models of cognitive phenotypes showed significant sex differences (χ²[df] = 127.5[21], P<0.001) in Clusters 2, females showed a higher proportion of combined H atrophy and ChP enlargement, while in Cluster 3, males exhibited a more diffuse distribution of structural patterns (Supplementary Figure 13, Supplementary Table S19). Finally, the association between CSF biomarkers and structural *N* measures did not differ significantly by sex (Supplementary Figure 14, Supplementary Table S20).

## Discussion

The present study demonstrates a substantial dissociation between amyloid positivity (*A*^+^) and structural neurodegeneration (*N*^+^) across the AD spectrum. Although H and IPL atrophy progressed at the group level and ChP enlargement increased with clinical severity, nearly half of amyloid-positive individuals exhibited no detectable macroscopic structural neurodegeneration. Consequently, conventional structural MRI markers showed limited utility for individual-level classification despite robust differences between diagnostic groups. These findings indicate that amyloid positivity (*A*^+^) alone does not uniformly predict downstream structural neurodegeneration (*N*^+^) and reveal considerably greater biological heterogeneity than implied by a simple linear model of disease progression.

This heterogeneity has important implications for our understanding of AD. Rather than representing a single, homogeneous clinicopathological entity, amyloid-positive (*A*^+^) AD may encompass multiple biologically distinct structural phenotypes that converge on a common molecular hallmark while differing in downstream neurodegenerative trajectories. We refer to these phenotypes collectively as amyloidopathies (Figure 4), analogous to the biological diversity recognised within tauopathies, synucleinopathies and TDP-43 proteinopathies. Importantly, our findings should not be interpreted as evidence against the central role of amyloid pathology in AD. Instead, they suggest that amyloid positivity (*A*^+^) alone incompletely captures the biological complexity of downstream neurodegeneration (*N*^+^). This interpretation is consistent with contemporary models demonstrating that brain ageing follows multiple interacting structural and biological trajectories rather than a single uniform pathway [31].

**Figure 4.**
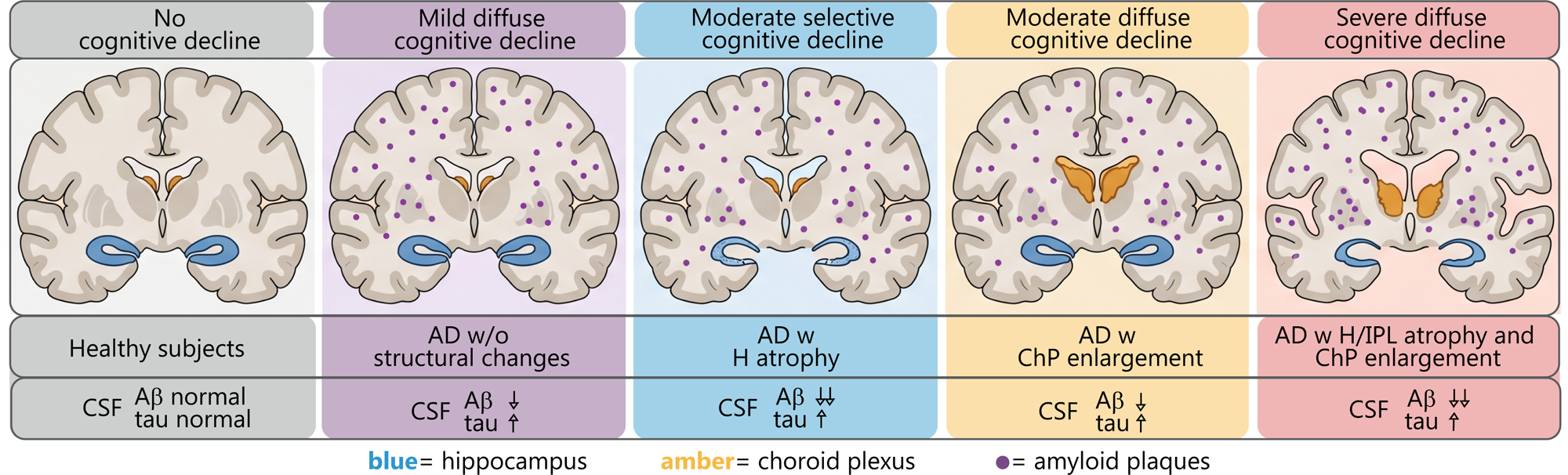
Spectrum of cognitive, structural, and biochemical alterations in amyloid-positive Alzheimer’s disease cases. Coronal brain sections illustrate four phenotypic subtypes characterised by distinct patterns of cognitive impairment, regional brain atrophy, and cerebrospinal fluid (CSF) biomarker changes. **Abbreviations**: Aβ, amyloid beta; ChP, choroid plexus; H, hippocampus; IPL, inferior parietal lobule; CSF, cerebrospinal fluid.

Several observations further support this interpretation. First, H and IPL atrophy were associated with significantly lower CSF Aβ_1–42_ concentrations, whereas ChP enlargement showed no such relationship. This suggests that distinct structural phenotypes may arise through partially different biological mechanisms. ChP enlargement may reflect alterations in cerebrospinal fluid dynamics, glymphatic clearance, blood-brain barrier integrity or neuroinflammatory processes rather than directly mirroring parenchymal amyloid deposition. Second, cognitive phenotypes mapped closely onto distinct structural signatures despite similar amyloid status, indicating that structural heterogeneity is clinically meaningful rather than simply reflecting measurement variability. Finally, the persistence of the amyloid-positive (*A*^+^)/neurodegeneration-negative (*N*^-^) phenotype across both sexes supports the robustness of this biological dissociation, although regional structural correlates of cognition demonstrated modest sex-related differences.

These findings also have important implications for biomarker-guided therapeutic strategies. Current anti-amyloid therapies are prescribed primarily on the basis of amyloid positivity (*A*^+^). Our data suggest that amyloid-positive (*A*^+^) individuals represent a biologically heterogeneous population with distinct structural phenotypes that may differ in disease stage, downstream pathophysiology or therapeutic responsiveness. Although our study was not designed to evaluate treatment response, the observed heterogeneity raises the possibility that structural phenotyping could complement molecular biomarkers when stratifying patients for future clinical trials and precision therapeutic approaches. Prospective studies incorporating longitudinal imaging and treatment outcomes will be required to determine whether these structural phenotypes predict differential responses to disease-modifying therapies.

In conclusion, our findings indicate that amyloid positivity (*A*^+^) and structural neurodegeneration (*N*^+^) are only partially coupled at the individual level. Recognising this heterogeneity provides a more nuanced framework for understanding AD biology and supports the development of biomarker-informed precision medicine approaches that integrate molecular, structural and cognitive phenotypes rather than relying on amyloid status alone.

## Limitations

Several limitations should be considered when interpreting these findings. First, the primary analyses were cross-sectional and therefore cannot establish the temporal sequence linking amyloid deposition, tau pathology, structural neurodegeneration, and cognitive decline. Although our longitudinal analyses demonstrated relative stability of structural phenotypes over four years, future studies incorporating repeated multimodal biomarker assessments will be required to define their biological trajectories more precisely.

Second, while the overall cohort was large (n = 872), subdivision into individual structural phenotypes resulted in relatively small subgroup sizes, reducing statistical power for some secondary analyses. Replication in larger independent cohorts will therefore be important to confirm the prevalence and biological characteristics of the proposed structural phenotypes.

Third, neurodegeneration was defined using macroscopic volumetric MRI, consistent with current clinical practice and the ATN framework. However, this approach is unlikely to detect earlier microstructural, synaptic, metabolic, or functional changes that may precede measurable tissue loss. Consequently, participants classified as structurally normal (N^−^) should not be interpreted as lacking neurodegeneration altogether, but rather as lacking detectable macroscopic structural abnormalities. Likewise, PET and CSF biomarkers provide indirect measures of amyloid and tau pathology and cannot fully capture regional, cellular, or temporal aspects of protein aggregation.

Fourth, we did not systematically assess the contribution of common age-related co-pathologies, including cerebrovascular disease, TDP-43 pathology, or α-synuclein deposition. These pathologies frequently coexist with amyloid pathology in older adults and may contribute to the structural and cognitive heterogeneity observed in our cohort.

Finally, participants were recruited from the ADNI, a research cohort comprising predominantly well-educated and highly motivated volunteers. Consequently, the generalisability of these findings to more diverse clinical populations with greater medical comorbidity, broader socioeconomic backgrounds, and different ethnic representation requires further validation.

Despite these limitations, the consistency of our findings across multiple structural biomarkers, cognitive phenotypes, machine-learning approaches, longitudinal analyses, and sex-stratified cohorts supports the robustness of the central observation that amyloid positivity and MRI-defined structural neurodegeneration are only partially coupled at the individual level.

## Conclusions

Our findings demonstrate substantial heterogeneity in the relationship between amyloid pathology, structural neurodegeneration, and cognitive impairment across the AD spectrum. Although robust group-level structural differences were observed, conventional MRI markers showed limited utility for predicting amyloid positivity (*A*^+^) at the individual level, with nearly half of amyloid-positive participants exhibiting no detectable macroscopic structural neurodegeneration.

These findings indicate that amyloid positivity (*A*^+^) alone does not uniformly predict downstream structural neurodegeneration and support the concept that AD comprises multiple biologically distinct structural phenotypes, collectively termed amyloidopathies. Rather than serving as standalone indicators of disease state, structural MRI measures should be interpreted alongside molecular and functional biomarkers to capture the full biological complexity of AD.

Recognising this heterogeneity has important implications for biomarker interpretation, patient stratification, clinical trial design, and the development of precision medicine approaches. Future longitudinal, multimodal, and treatment-response studies will be essential to determine how these distinct structural phenotypes arise and whether they predict differential clinical progression and therapeutic response.

## Statements and Declarations

### Ethics statement

The authors confirm that all procedures contributing to this work comply with the ethical standards of the relevant national and institutional committees on human experimentation and with the Helsinki Declaration of 1975, as revised in 2008. The study adhered to the Strengthening the Reporting of Observational Studies in Epidemiology (STROBE) guidelines for cross-sectional studies. Written informed consent was obtained from all participants or heir authorized representatives at the time of enrollment in the Alzheimer’s Disease neuroimaging Initiative (ADNI).

### Authors contribution

GBS: Conceptualization, Methodology, Supervision, Funding acquisition, Writing – original draft, Writing – review & editing.

JSN: Data curation, Formal analysis, Visualization, Software, Writing – original draft, Writing – review & editing.

MČ: Validation, Interpretation of results, Writing – review & editing.

LL: Validation, Interpretation of results, Writing – review & editing.

TK: Validation, Interpretation of results, Writing – review & editing.

All authors have read and approved the final version of the manuscript.

### Data availability

The dataset analyzed for this study are publicly available through the Alzheimer’s Disease neuroimaging Initiative (ADNI) database (https://adni.loni.usc.edu/). Access is granted upon registration and institutional approval. No new primary data were generated for this study; all findings are based on secondary analysis of ADNI repository.

## Supplementary Figure Legends

**Supplementary Figure 1. Group-level associations between brain region volumes and diagnostic status.**

Multinomial logistic regression-derived odds ratios (95% CI) for diagnostic group membership (MCI or AD dementia vs. healthy subjects) per 0.1-unit decrease (H/IPL) or increase (ChP) in normalised regional volume.

**Supplementary Figure 2. Individual-level diagnostic performance of machine learning predictions based on isolated regional volumes**.

Receiver operating characteristic (ROC) curves of multiclass diagnostic predictions based on H, IPL, or ChP volumes, computed using (**a**) Random Forest, and (**b**) multinomial logistic regression.

**Abbreviations**: HS: healthy subjects, MCI: AD-related mild cognitive impairment, D: Alzheimer’s dementia, H: hippocampus, IPL: Inferior parietal lobule, ChP: choroid plexus.

**Supplementary Figure 3. Individual-level diagnostic performance of machine learning predictions based on combined regional volumes**.

Receiver operating characteristic (ROC) curves of multiclass diagnostic predictions based on various ROI combinations, computed using (**a**) artificial neural network, (**b**) Random Forest, and (**c**) multinomial logistic regression.

**Abbreviations**: HS: healthy subjects, MCI: AD-related mild cognitive impairment, D: Alzheimer’s dementia, H: hippocampus, IPL: inferior parietal lobule, ChP: choroid plexus.

**Supplementary Figure 4. Distribution of regional brain volumes across diagnostic groups**.

Scatter plots illustrating individual variability in normalized brain volumes (H, IPL, and ChP) stratified by diagnostic category.

**Abbreviations**: HS: healthy subjects, MCI: AD-related mild cognitive impairment, D: Alzheimer’s dementia.

**Supplementary Figure 5. Distribution of APOE 4 alleles across volumetric phenotypes**.

Bar charts showing the prevalence and frequency of APOE ε4 alleles stratified by specific patterns of volumetric abnormalities in the MCI and AD dementia cohorts.

**Supplementary Figure 6. Machine-learning-based diagnostic prediction using categorical structural abnormalities**.

Receiver operating characteristic (ROC) curves showing multiclass diagnostic predictions generated by artificial neural network models using categorical structural abnormalities (presence of atrophy or enlargement). Models were based on (**a**) isolated regional abnormalities and (**b**) combined categorical abnormality profiles.

**Supplementary Figure 7. Global and domain-specific cognitive performance by volumetric phenotype**.

(**a**) Mini-Mental State Examination (MMSE) total scores stratified by volumetric phenotype and diagnostic group.

(**b-e**) ADNI CogSum domain scores (Memory, Executive Function, Language, and Visuospatial) stratified by volumetric phenotype and diagnostic group.

**Abbreviations**: HS: healthy subjects, MCI: AD-related mild cognitive impairment, D: Alzheimer’s dementia, H: hippocampus, IPL: inferior parietal lobule, ChP: choroid plexus.

**Supplementary Figure 8. Cerebrospinal fluid (CSF) biomarker concentrations by volumetric phenotype**.

Comparison of (**a**) CSF Aβ_1-42_, (**b**) total tau, and (**c**) phosphorylated tau (p-tau181) levels stratified by the presence or absence of specific regional volumetric abnormalities.

*P < 0.05, **P < 0.01.

**Supplementary Figure 9. Sex-specific associations between structural brain changes (*N*) and diagnostic status.**

Sex-specific distributions of normalised H, IPL, and ChP volumes across HS (*A*^-^) and amyloid-positive (*A*^+^) participants with MCI or D.

**Abbreviations**: HS, healthy subjects; MCI, AD-related mild cognitive impairment; D, Alzheimer’s dementia; H, hippocampus; IPL, inferior parietal lobule; ChP, choroid plexus.

**Supplementary Figure 10. Sex-specific prevalence of structural neurodegeneration (*N*) in A^+^ individuals**.

Venn diagrams illustrating overlap between regional structural abnormalities and the proportion of *A*^+^ cases with no detectable structural abnormalities (*N*^-^) in (**a**) females and (**b**) males.

**Supplementary Figure 11. Sex-specific individual-level diagnostic performance of artificial neural network predictions based on regional volumes**.

Comparison of receiver operating characteristic (ROC) curves fr multiclass diagnostic predictions based on H, IPL, or ChP volumes between sexes, using (**a**) individual regional volumes and (**b**) combined regional volumes.

**Abbreviations**: HS, healthy subjects; MCI, AD-related mild cognitive impairment; D, Alzheimer’s dementia; H, hippocampus; IPL, inferior parietal lobule; ChP, choroid plexus.

**Supplementary Figure 12. Sex-specific individual-level diagnostic performance of artificial neural network predictions based on categorical structural abnormalities**.

Comparison of receiver operating characteristic (ROC) curves for multiclass diagnostic predictions based on the presence of atrophy or enlargement between sexes, using (**a**) individual structural abnormalities and (**b**) combined structural abnormality profiles.

**Abbreviations**: HS, healthy subjects; MCI, AD-related mild cognitive impairment; D, Alzheimer’s dementia; H, hippocampus; IPL, inferior parietal lobule; ChP, choroid plexus.

**Supplementary Figure 13. Sex-specific differences in structural phenotypes based on cognitive profiles**.

Sex-specific distribution of volumetric phenotypes within each cognitive cluster (row-wise percentages).

**Abbreviations**: N, no volumetric change; H, hippocampal atrophy; P, inferior parietal lobule atrophy; C, choroid plexus enlargement; HP, hippocampal and inferior parietal lobule atrophy; HC, hippocampal atrophy with choroid plexus enlargement; HPC, hippocampal and inferior parietal lobule atrophy with choroid plexus enlargement.

**Supplementary Figure 14. Sex-specific differences in cerebrospinal fluid (CSF) biomarker concentrations across volumetric phenotypes**.

Sex-related differences in (**a**) CSF Aβ_1-42_, (**b**) total tau, and (**c**) phosphorylated tau (p-tau181) concentrations stratified by the presence or absence of specific regional volumetric abnormalities.

