## Supplementary figures and images for "Structural heterogeneity reveals distinct amyloidopathies in Alzheimer’s disease"

### Supplementary Figure 1

#### Supplementary Table 1

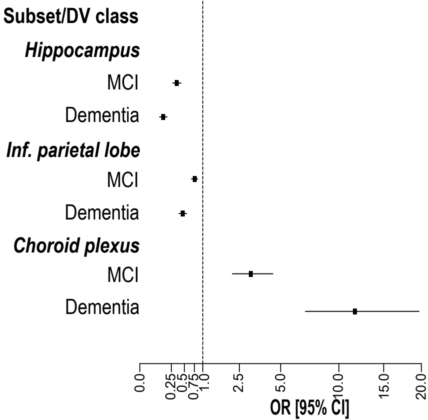

### Supplementary Figure 2

#### Supplementary Table 2

a

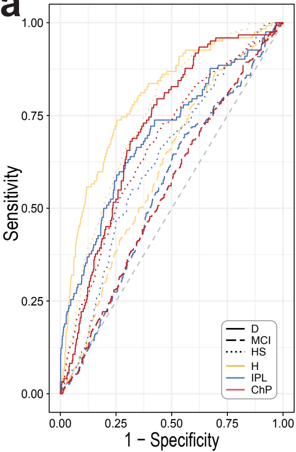


b

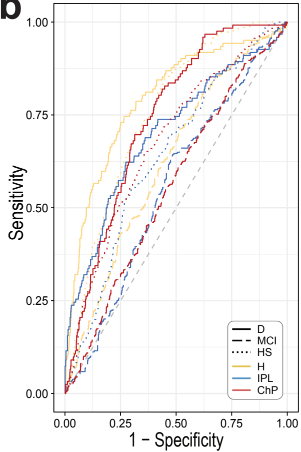

### Supplementary Figure 3

#### Supplementary Table 3

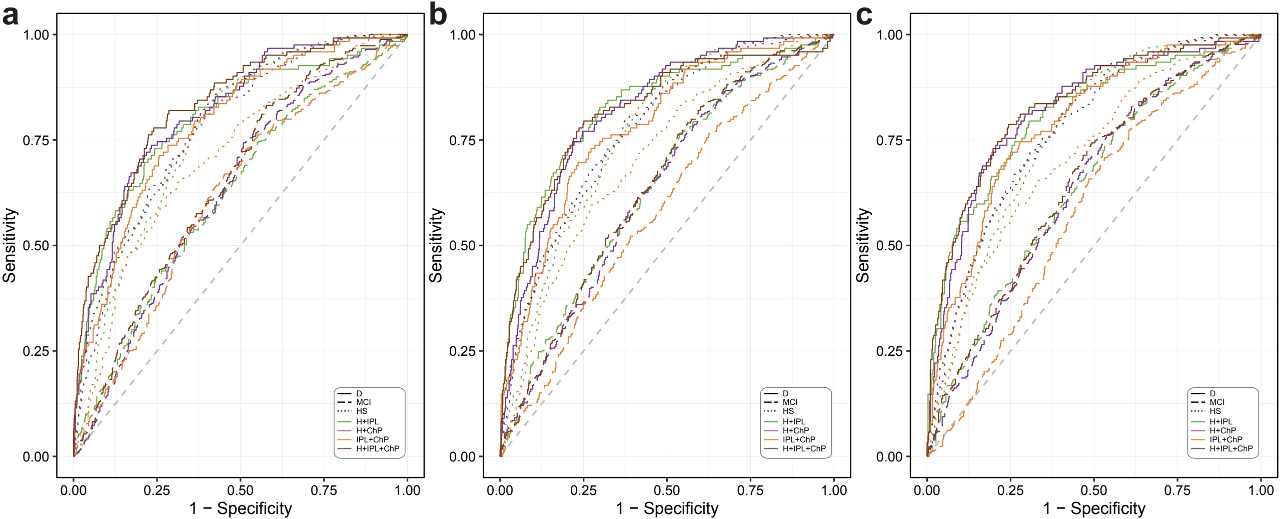

### Supplementary Figure 4

#### Supplementary Table 4

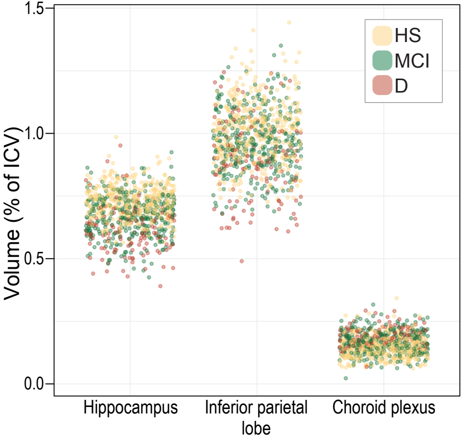

### Supplementary Figure 5

#### Supplementary Table 5

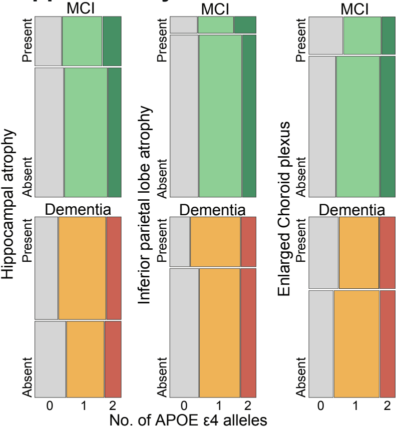

### Supplementary Figure 6

#### Supplementary Table 6

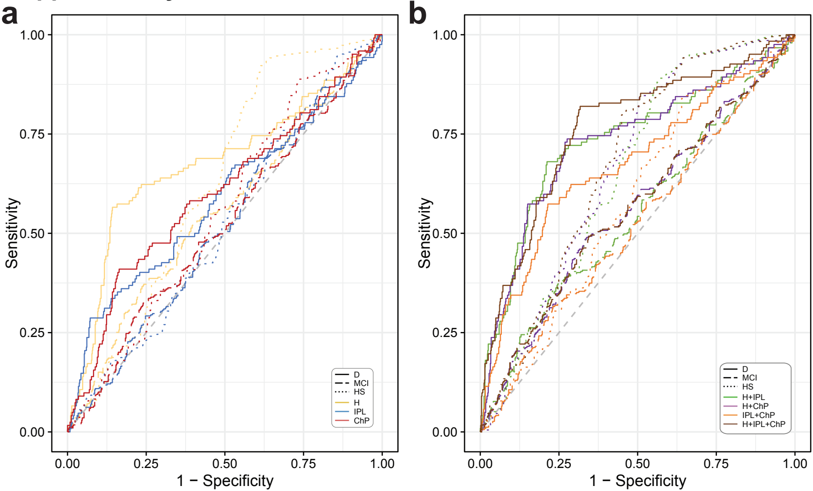

### Supplementary Figure 7

#### Supplementary Table 7

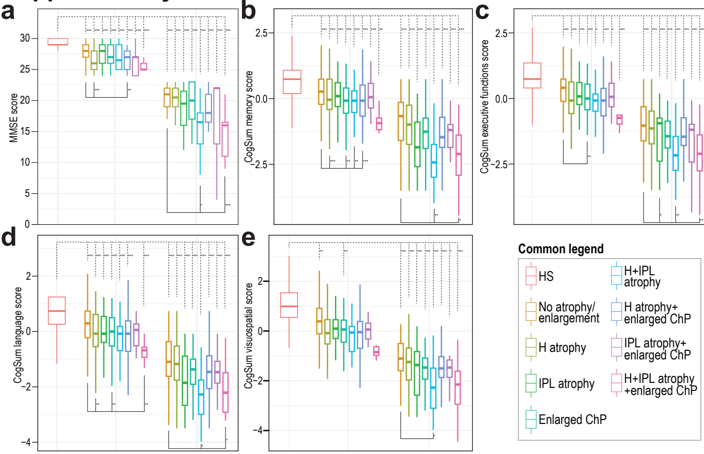

### Supplementary Figure 8

#### Supplementary Table 8

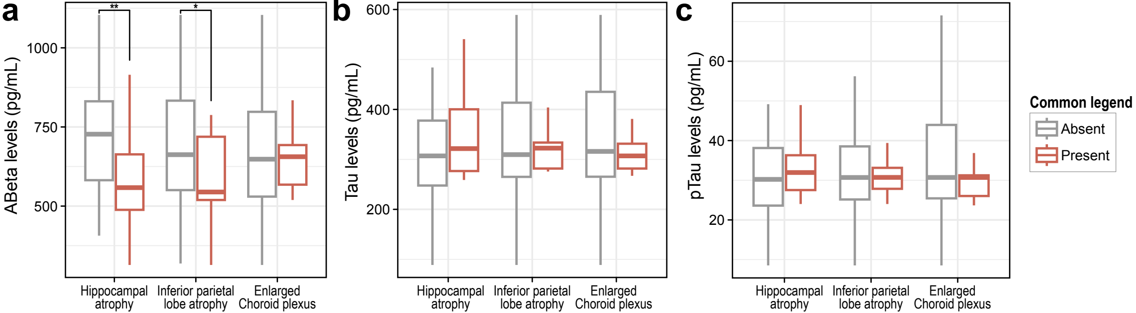

### Supplementary Figure 9

#### Supplementary Table 9

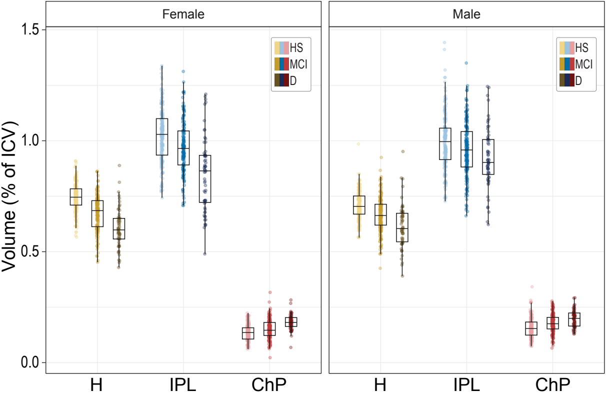

### Supplementary Figure 10

#### Supplementary Table 10

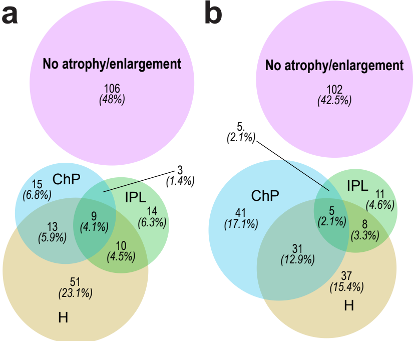

### Supplementary Figure 11

#### Supplementary Table 11

a

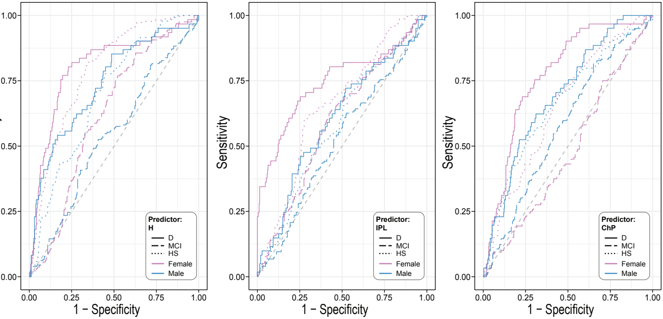


b

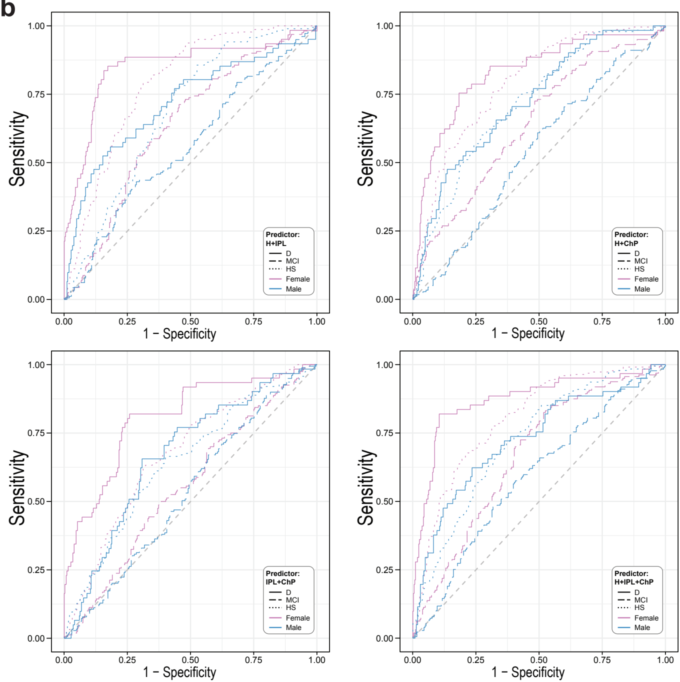

### Supplementary Figure 12

#### Supplementary Table 12

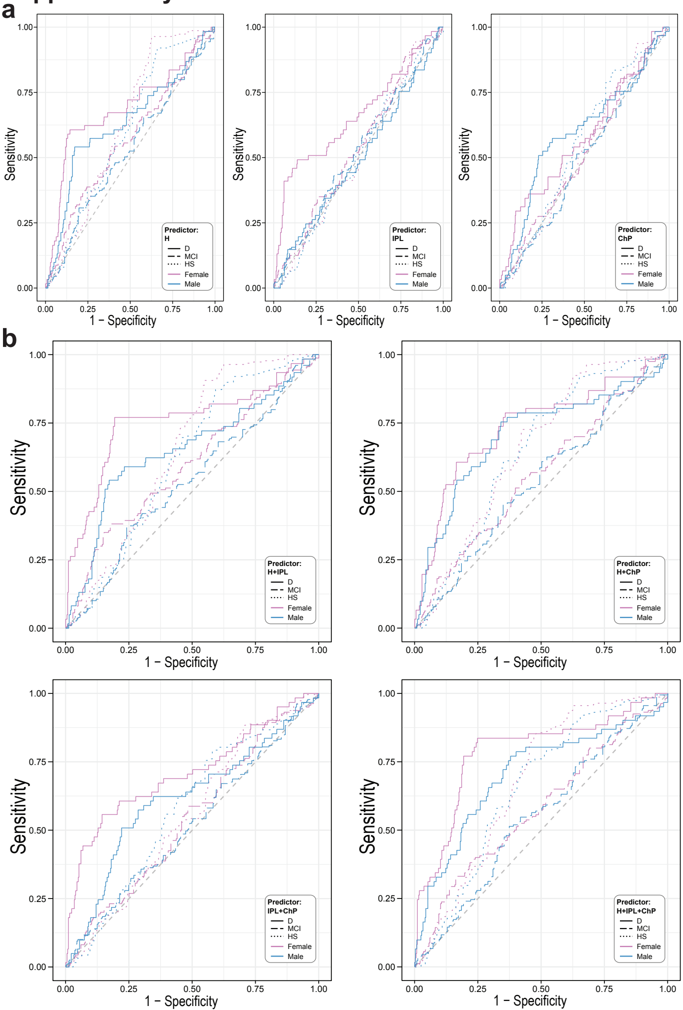

### Supplementary Figure 13

#### Supplementary Table 13

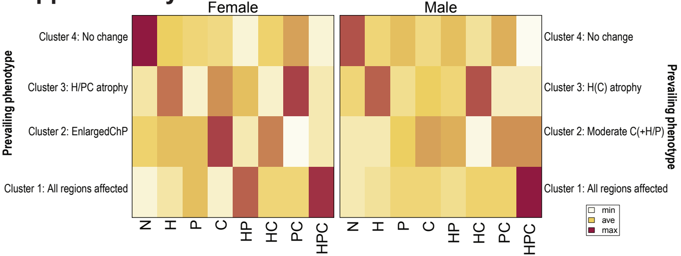

### Supplementary Figure 14

#### Supplementary Table 14

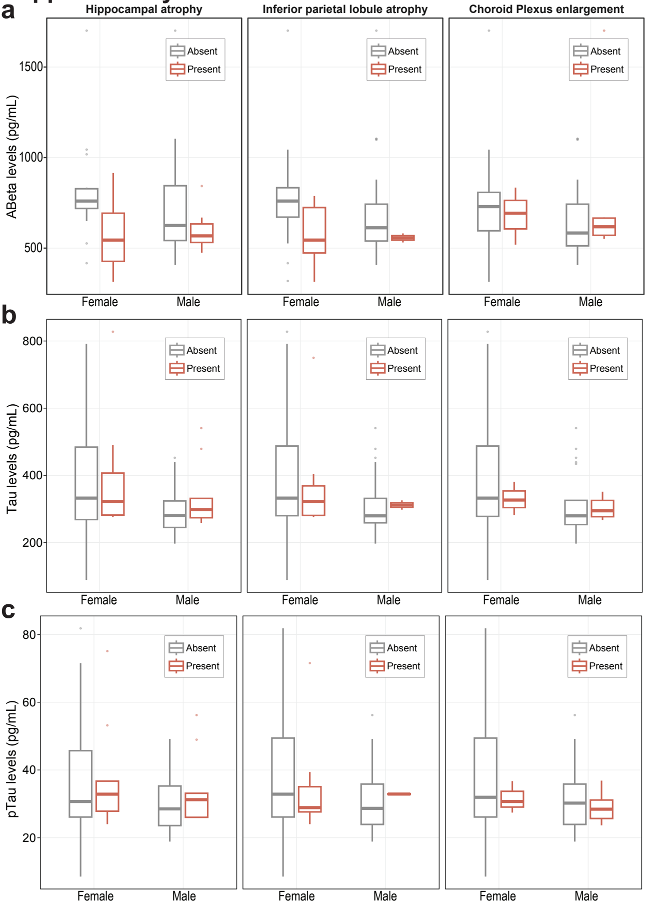
